# Duration of intestinal mucosal antibody responses to poliovirus in children routinely immunised with bivalent oral polio vaccine and inactivated polio vaccine in Tanzania: A longitudinal cohort and cross-sectional study

**DOI:** 10.64898/2026.05.29.26354450

**Authors:** Alexandra A. Cordeiro, Naomi Miall, Myung Hee Lee, Margaret E. Ackerman, Joshua A. Weiner, Samia Sami, Onike Mcharo, Enock Miyaye, Loyce Mhango, Nsia Ulomi, Audrey Godin, Minetaro Arita, Rachel M. Burke, Oraine B. L.Campbell, Danielle de Jong, Pytsje T. Hoekstra, Govert J. van Dam, Paul Corstjens, Maureen Ward, Lars F. Westblade, Adolfine Hokororo, Safari Kinung’hi, Wendy Wieland-Alter, Ruth I. Connor, Elizabeth B. Brickley, Jennifer A. Downs, Peter F. Wright

**Author notes:** Co-first author (Alexandra A. Cordeiro, Naomi Miall). Co-senior author (Elizabeth B. Brickley, Jennifer A. Downs, Peter F. Wright).

## Abstract

**Background:** Mucosal immunity is critical for preventing poliovirus transmission. Despite evidence that infant immunisation protects against poliovirus infection into adulthood, the duration of vaccine-induced intestinal antibody responses remains poorly characterised.

**Methods:** We evaluated poliovirus type-specific neutralising activity and immunoglobulin levels in stool and serum from children in Tanzania who completed routine poliovirus vaccine series (bivalent oral polio vaccine at birth, 6, 10, and 14-weeks, and inactivated polio vaccine at 14-weeks). The study included a longitudinal cohort with four visits over 6 months and a cross-sectional sample of children recruited 1 to 108-months after vaccine series completion. Potential modification by nutritional factors, gastrointestinal infections, and environmental enteropathy was explored.

**Results:** Among 103 longitudinal and 246 cross-sectional participants enrolled, 33% and 18% had positive poliovirus type-1 (PV1) stool neutralisation, and 66% and 56% had positive poliovirus type-3 (PV3) neutralisation 1 month after vaccination. All were seropositive for PV1 and PV3 across timepoints. Infants followed longitudinally who were stool neutralisation-positive at enrolment had no boost in neutralisation after vaccination, while those stool neutralisation-negative at enrolment experienced a weak boost at 1 month. Stool neutralisation half-life among longitudinal cohort infants was 3.4 months [95% CI 2.6-5.0] for PV1 and 1.7 months [1.4-2.3] for PV3. Moderate evidence suggested concurrent viral intestinal infections were associated with lower neutralisation responses (PV1 p=0.153; PV3 p=0.052).

**Conclusion:** Intestinal antibody responses to poliovirus vaccination were short-lived. The impact of waning intestinal antibodies on transmission risk remains unclear and research is needed to identify vaccination strategies that induce durable mucosal immunity.

## Introduction

Oral polio vaccines (OPV) induce primary intestinal mucosal immunity that inhibits enteric poliovirus (PV) replication and reduces the risk of onward transmission, while also generating systemic immunity that protects individuals against paralytic poliomyelitis disease.^1^ Inactivated polio vaccine (IPV) primarily protects against poliomyelitis by inducing robust serum antibody responses, and is also capable of boosting intestinal mucosal immunity in individuals with previous live PV exposure.^2^

While previous research suggests PV-specific serum antibodies persist for at least 15-years after vaccination,^3^ the duration of vaccine-induced intestinal mucosal antibodies remains to be established, with existing evidence primarily based on trials with two-months or less of follow-up.^4–7^ Moreover, it remains uncertain how co-factors modify the induction and kinetics of intestinal antibody responses. It is well documented that oral vaccines, including OPV, demonstrate lower effectiveness among children in low- and middle-income country settings than in high-income countries.^8, 9^ It is hypothesised that factors such as transplacental and breastmilk-derived maternal antibodies, concurrent enteric pathogen exposure, and environmental enteropathy (EE) may reduce oral vaccine immunogenicity.^8^

This study aims to evaluate the duration and determinants of intestinal antibody responses to PV vaccines delivered through routine immunisation in rural northwestern Tanzania, where children receive four doses of bivalent OPV (bOPV, containing poliovirus types 1 and 3 [PV1 and PV3]) at birth, 6, 10, and 14-weeks of age and one dose of IPV (containing PV serotypes 1, 2 and 3) at 14-weeks. PV type-specific neutralisation and immunoglobulin A (IgA) were quantified in stool, and PV type-specific neutralisation, IgA and immunoglobulin G (IgG) levels were quantified in serum, collected from a cohort followed longitudinally for 6-months and a cross-sectional study of children enrolled one-month to 9 years after completing the primary polio vaccine series. The study objectives were to evaluate the duration and trajectory of PV type-specific intestinal antibodies over childhood and to evaluate their associations with serum antibody responses, nutritional status, and exposure to concomitant gastrointestinal infections.

## Methods

### Study design, settings, and participants

The study was conducted at two community health clinics in Ilemela District, northwestern Tanzania (Supplementary Figure 1). Ilemela District is located in a schistosomiasis endemic region with a documented *Schistosoma mansoni* prevalence of >80% among children aged 1-5 years.^10^ Eligible children had: (1) documented completion of the four-dose bOPV and single IPV vaccine series; (2) written informed consent from a parent/guardian aged ≥18-years and (3) a mother willing to undergo HIV-1 rapid testing or to provide HIV-1 status documentation.

Between November 2023 and November 2025, children were enrolled and stratified into two groups according to time since receipt of the final bOPV dose administered at 14-weeks of age. Infants within 7-days of completing the vaccine series were enrolled into a longitudinal cohort and evaluated at enrolment and 1, 3, and 6-months post-enrolment. Infants and children who had previously completed the vaccine series were enrolled into a cross-sectional study, evaluated at a single visit, and grouped according to time since vaccine series completion: 1, 3, 6, 12, 24, 36, and 96 to 108-months (8 to 9-years) (Figure 1). Children in the oldest stratum (aged 8-9 years at enrolment) did not receive IPV, which was first introduced to the routine immunisation schedule in 2018, and received trivalent OPV (tOPV) instead of bOPV. A sub-national novel OPV type-2 (nOPV2) campaign was conducted in Tanzania in 2023, however the study region was not included.

**Figure 1.**
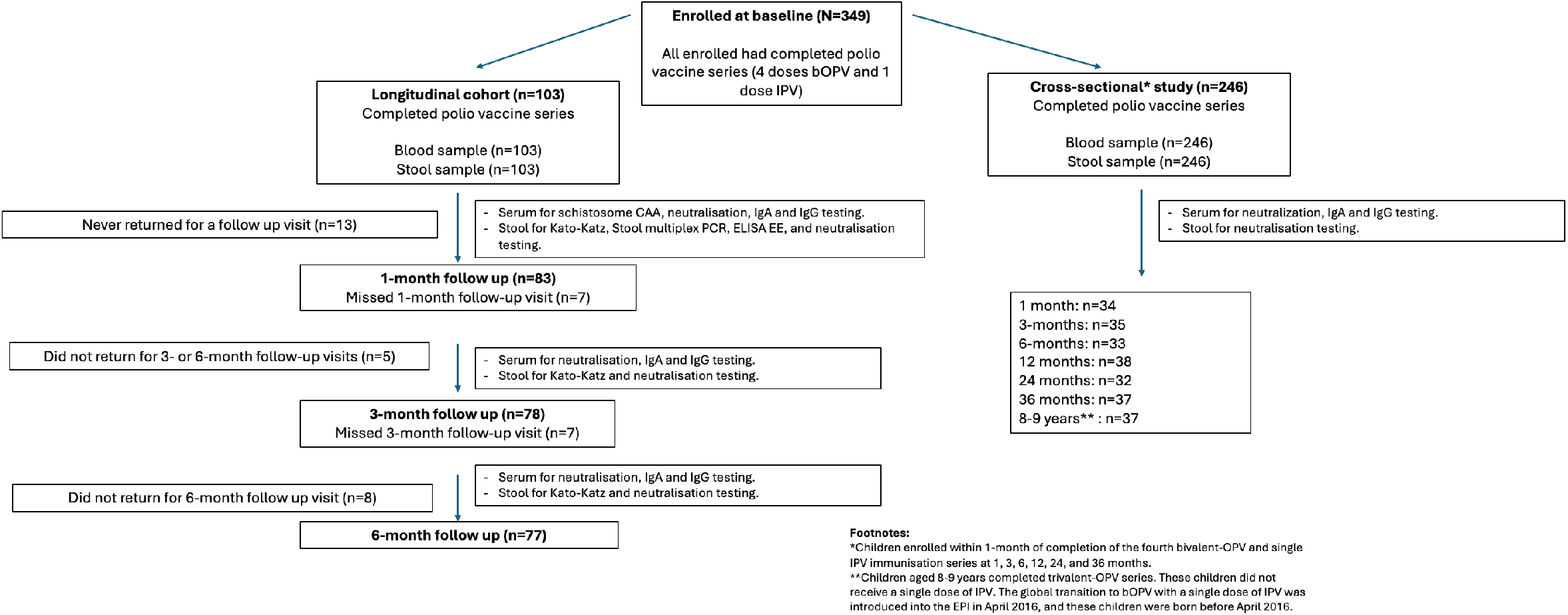
Flow diagram of study recruitment and participant selection for children in the longitudinal cohort and cross-sectional study in northwestern Tanzania (November 2023-May 2025). Infants were enrolled into the longitudinal cohort within 7 days after polio vaccine series completion and followed prospectively for 6-months with stool and blood samples collected at each visit. Children in the cross-sectional study were enrolled at pre-defined time points (1, 3, 6, 12, 24, 36, and 8-9 years) after completion of the same polio vaccine series with a single stool and blood sample collected at enrolment. Eligibility for children in the cross-sectional study was defined by time-since-vaccination, allowing enrolment up to one-month before and after the target interval (i.e., up to 13 months for the 12 month group.

Parent(s)/guardians provided written informed consent for child participation. Children >7-years subsequently provided written assent. Ethical approvals were obtained from the National Health Research Ethics Committee in Tanzania (NIMR/HQ/R.8a/Vol.IX/4390), Weill Cornell Institutional Review Board (#22-12025526) and Dartmouth-Hitchcock Institutional Review Board (MOD00019059). Free treatment was provided for any infections diagnosed during the study period, as clinically indicated.

### Data and sample collection

At enrolment, trained study nurses measured participants’ anthropometrics and collected basic sociodemographic information and clinical history, including age, sex, breastfeeding status, and history of systemic or gastrointestinal illness from parents/guardians using an orally administered questionnaire in Kiswahili (English translation available in Supplementary Materials). For infants in the longitudinal cohort, parents/guardians also reported socioeconomic indicators derived from the Oxford Multidimensional Poverty Index including parental education, household ownership of assets, and access to electricity, clean water and sanitation facilities. Routine poliovirus and rotavirus immunisation histories were abstracted from participants’ vaccination cards, supplemented by caregiver-reported receipt of poliovirus vaccines during supplementary immunisation activities in their communities. At enrolment, children whose mothers were living with HIV underwent HIV-1 rapid testing if aged ≥24-months, or provided dried blood spots for HIV-1 RNA quantification at Bugando Medical Center (Mwanza City, TZA) if aged<24-months. Stool (~4-5 g) and venous blood (2 mL) were collected at every visit. For infants in the longitudinal cohort, 4g of stool collected at enrolment was diluted in 4mL of Cary Blair transport media for subsequent multiplex PCR testing. All samples were transported to the National Institute for Medical Research (NIMR) laboratory (Mwanza, TZA) and stored at −80°C, with aliquots shipped on dry ice to Weill Cornell Medicine (New York, USA) and Dartmouth College (New Hampshire, USA).

### Laboratory Procedures

At the study health clinics, trained parasitologists examined stool Kato-Katz slides microscopically for presence of parasite eggs within 1 hour of collection due to egg fragility, and after 24-hours for confirmation.^11^. At the NIMR laboratory, serum schistosome circulating anodic antigen (CAA) was quantified using up-converting reporter particle technology.^12^ CAA concentrations ≥30 pg/mL were considered positive. EE analyses were performed on stool samples collected at enrolment for infants in the longitudinal cohort using enzyme-linked immunosorbent assays (ELISAs) to quantify indicators of intestinal inflammation (i.e., neopterin (Aviva, USA) and myeloperoxidase (Aviva, USA)) and intestinal permeability (i.e., alpha-1 antitrypsin (Immundiagnostik, Germany)).^13^ ELISAs were performed according to manufacturers’ instructions, except for neopterin, for which stool samples were diluted 1:5 in 0.9% NaCl.^14^ At Weill Cornell Medicine, stool samples were tested for 22 different gastrointestinal pathogens using a multiplex PCR BIOFIRE FILMARRAY Gastrointestinal panel (bioMérieux, Salt Lake City, UT, USA).^15^

Poliovirus types 1, 2, and 3 (PV1, PV2, and PV3)-specific neutralising and binding antibody responses were measured in all stool and serum samples. Neutralisation titres were reported as the reciprocal of the dilution needed to achieve 60% neutralisation of luciferase-expressing polio pseudo-viruses.^16^ Titres <4 (i.e., the lower limit of detection) were recorded as 2 and titres ≥8 were considered positive. PV serotype-specific IgA in stool and IgA and IgG in serum were quantified by a multiplex assay using monovalent inactivated PV vaccines coupled to fluorescently-coded magnetic microspheres, and expressed as median fluorescence intensity (MFI).^5^ Values that were negative after background signal subtraction were recorded as half of the lowest observed homotypic MFI. Stool samples were tested at a dilution of 1:100, serum samples were tested at 1:250 for IgA and 1:1000 for IgG. Pooled human serum–derived IgA and IgG were used as positive controls for each plate. Total stool IgA (μg/mL) was measured using the same multiplex assay.

### Statistical analysis

Enroling 100 infants into the longitudinal cohort provided >80% power to detect fold-changes ≥2 in stool and serum antibody neutralisation titres between time points on the log-scale, assuming a within-subject log-titre standard deviation (SD) ≤3.75 and ≤20% loss to follow-up. In the cross-sectional study, enroling 34 children per time point provided >84% power to detect a one-dilution step difference in neutralisation titres between time points on the log scale, assuming a SD of ≤1.

Analyses were performed using Stata (version 19/SE) and R (version 4.4.1). Descriptive statistics summarised sociodemographic characteristics. Stunting (i.e., low height-for-age, defined as a Z-score <2 SD below the World Health Organization (WHO) child growth standards median) was calculated for all participants <8 years of age.^17^ Exclusive breastfeeding at 4 months was inferred using caregiver-reported breastfeeding status at enrolment and the reported age at cessation of exclusive breastfeeding. Pathogens identified in stool by multiplex PCR were categorised by clinical relevance as enteroinvasive bacteria associated with dysentery, other gastrointestinal bacterial pathogens, viral infections, and *Giardia lamblia* (Supplementary Table 1). A composite EE score (range 0–10) was calculated from stool concentrations of neopterin, myeloperoxidase, and alpha-1 antitrypsin, with higher composite scores indicating greater EE severity.^13^ Scores in the upper quartile (≥7) were defined as high EE.

Median log_2_-transformed neutralising antibody titres and log_10_-transformed IgA and IgG MFI in stool and serum were summarisedd for each participant group and timepoint. Differences across timepoints were assessed using mixed-effects linear regression (for neutralisation titres, IgA and IgG MFI) or logistic regression (for positive neutralisation), with timepoint specified as a categorical variable to allow for non-linear changes over time, and individual participant fitted as a random-effect to account for repeated observations in the longitudinal cohort. Differences between PV1 and PV3 responses were assessed using mixed-effects linear regression models with participant-level random-effects for continuous neutralisation and an extension of McNemar’s test to account for repeated observations for the binary outcome of positive stool neutralising activity.^18^ Half-lives of stool neutralising antibodies were estimated in the longitudinal cohort using samples collected between 1 and 6 months post-vaccination using mixed-effects linear regression, with time since vaccination and age at vaccination as fixed-effects, and individual participant as a random effect. Logistic regression adjusted for age at vaccination, was used to assess factors associated with positive PV1 and PV3-specific stool neutralising activity at any timepoint in the longitudinal cohort. A t-test was used to compare log_2_-transformed serum neutralisation titers between participants with positive or negative homotypic stool neutralisation at enrolment. In sensitivity analyses, the outcome was redefined as detectable neutralising activity (titre ≥1:4) at any visit. Kendall’s tau-b correlations assessed associations between PV1 and PV3 stool and serum neutralisation, IgA, and IgG responses for longitudinal samples collected at the 1-month visit and all cross-sectional samples. Longitudinal participants with missing visits or incomplete samples (n=37) were included in the primary analyses and excluded in a sensitivity analysis.

## Results

A total of 103 infants were enrolled in the longitudinal cohort, and 246 children in the cross-sectional study from two lakeside communities in northwestern Tanzania (Figure 1, Supplementary Figure 1). Sociodemographic characteristics were similar between groups (Table 1). The median [IQR] age at receipt of the fourth bOPV dose was 16 (IQR: 15–17) weeks in both longitudinal and cross-sectional study groups. Most participants were exclusively breastfed at age 4 months (91% in the longitudinal cohort and 99% in the cross-sectional study), and approximately one-third were stunted at the time of enrolment (30% and 29% among the longitudinal and cross-sectional study participants younger than 8-years). Overall, 6% of participants were HIV-exposed but uninfected at birth, and one participant in the cross-sectional study was living with HIV at enrolment. Among infants in the longitudinal cohort, 14% tested positive for schistosome infection at enrolment with a median CAA concentration of 44.0 (IQR: 38.1–82.1) pg/mL, and 14% had *Entamoeba* cysts detected by stool microscopy. Multiplex stool PCR showed that 82% of infants in the longitudinal cohort were positive for at least one gastrointestinal infection, with a high prevalence of non-dysentery bacterial infections (71%) and viral infections (40%) (Table 1).

**Table 1.**
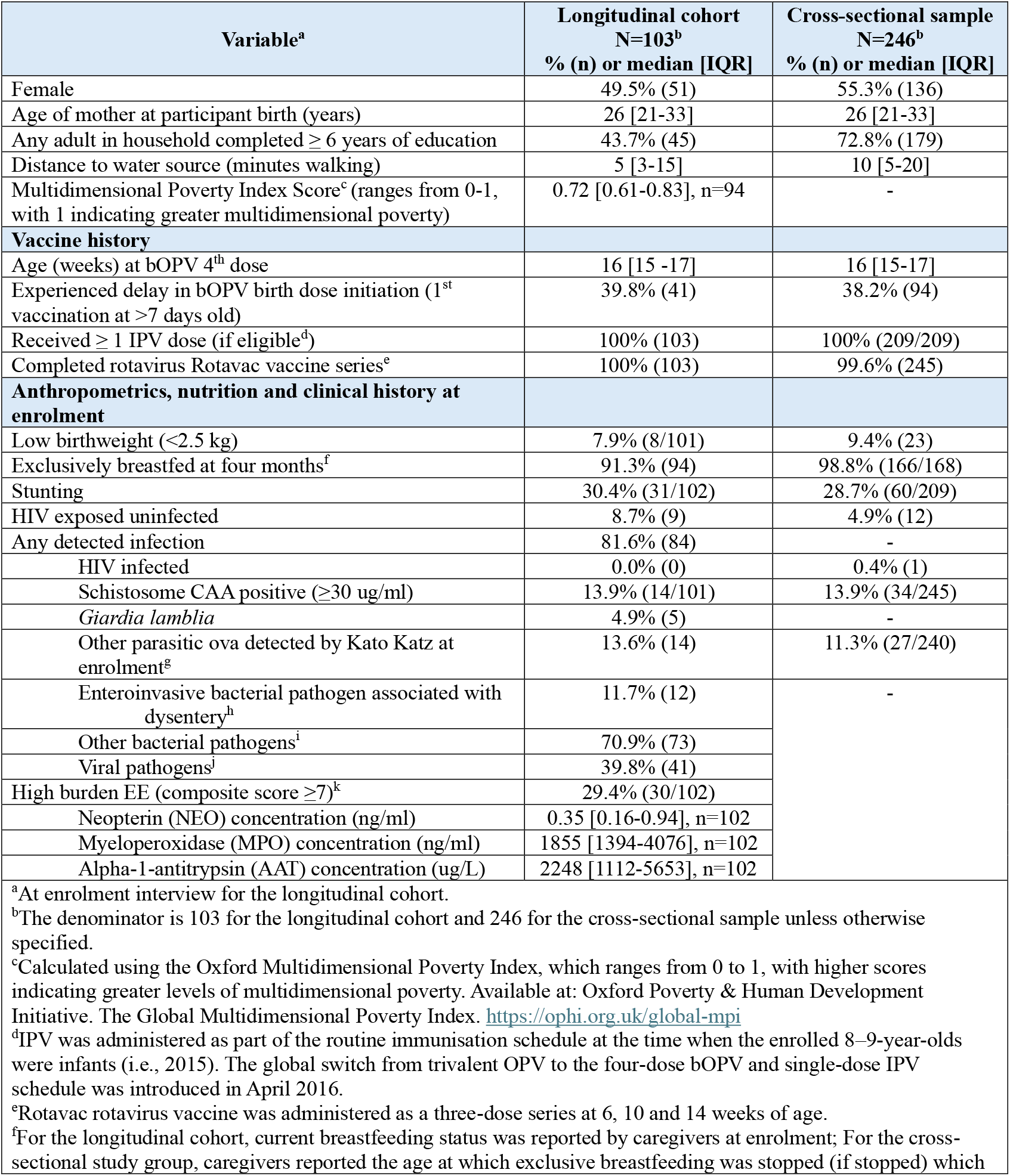
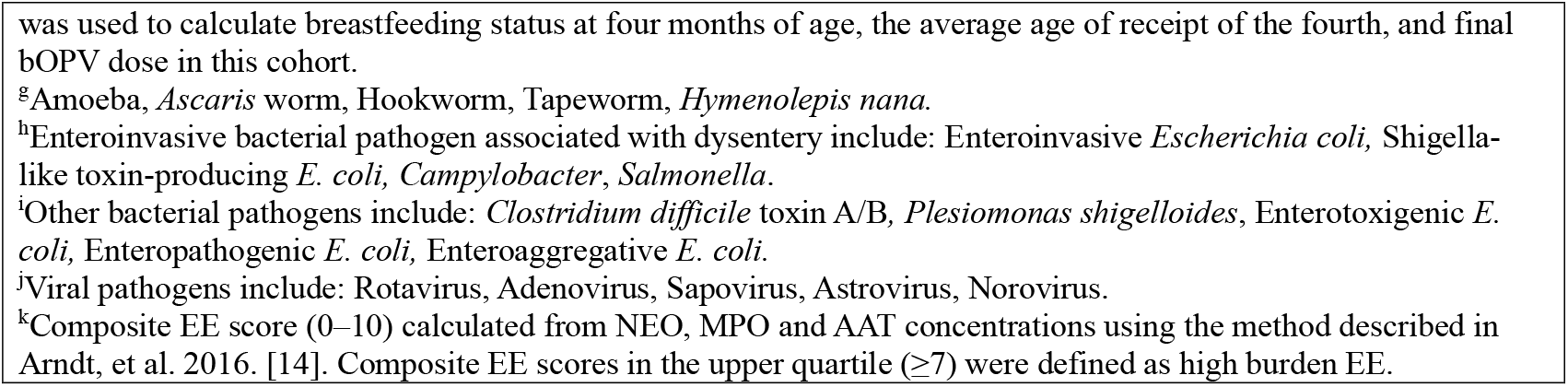
Sociodemographic characteristics, clinical history and laboratory diagnosis of infections measured at enrolment in 103 children recruited to the longitudinal cohort and 246 children recruited to the cross-sectional study. All participants had completed a routine four-dose bOPV and single-dose IPV vaccine series in Tanzania.

| Variable<sup>a</sup> | Longitudinal cohort<br>N=103<sup>b</sup><br>% (n) or median [IQR] | Cross-sectional sample<br>N=246<sup>b</sup><br>% (n) or median [IQR] |
| --- | --- | --- |
| Female | 49.5% (51) | 55.3% (136) |
| Age of mother at participant birth (years) | 26 [21-33] | 26 [21-33] |
| Any adult in household completed $\geq 6$ years of education | 43.7% (45) | 72.8% (179) |
| Distance to water source (minutes walking) | 5 [3-15] | 10 [5-20] |
| Multidimensional Poverty Index Score<sup>c</sup> (ranges from 0-1, with 1 indicating greater multidimensional poverty) | 0.72 [0.61-0.83], n=94 | - |
| **Vaccine history** |  |  |
| Age (weeks) at bOPV 4<sup>th</sup> dose | 16 [15 -17] | 16 [15-17] |
| Experienced delay in bOPV birth dose initiation (1<sup>st</sup> vaccination at >7 days old) | 39.8% (41) | 38.2% (94) |
| Received $\geq 1$ IPV dose (if eligible<sup>d</sup>) | 100% (103) | 100% (209/209) |
| Completed rotavirus Rotavac vaccine series<sup>e</sup> | 100% (103) | 99.6% (245) |
| **Anthropometrics, nutrition and clinical history at enrolment** |  |  |
| Low birthweight (<2.5 kg) | 7.9% (8/101) | 9.4% (23) |
| Exclusively breastfed at four months<sup>f</sup> | 91.3% (94) | 98.8% (166/168) |
| Stunting | 30.4% (31/102) | 28.7% (60/209) |
| HIV exposed uninfected | 8.7% (9) | 4.9% (12) |
| Any detected infection | 81.6% (84) | - |
| HIV infected | 0.0% (0) | 0.4% (1) |
| Schistosome CAA positive ($\geq 30$ ug/ml) | 13.9% (14/101) | 13.9% (34/245) |
| *Giardia lamblia* | 4.9% (5) | - |
| Other parasitic ova detected by Kato Katz at enrolment<sup>g</sup> | 13.6% (14) | 11.3% (27/240) |
| Enteroinvasive bacterial pathogen associated with dysentery<sup>h</sup> | 11.7% (12) | - |
| Other bacterial pathogens<sup>i</sup> | 70.9% (73) |  |
| Viral pathogens<sup>j</sup> | 39.8% (41) |  |
| High burden EE (composite score $\geq 7$)<sup>k</sup> | 29.4% (30/102) | |
| Neopterin (NEO) concentration (ng/ml) | 0.35 [0.16-0.94], n=102 |  |
| Myeloperoxidase (MPO) concentration (ng/ml) | 1855 [1394-4076], n=102 |  |
| Alpha-1-antitrypsin (AAT) concentration (ug/L) | 2248 [1112-5653], n=102 |  |

<sup>a</sup>At enrolment interview for the longitudinal cohort.
<sup>b</sup>The denominator is 103 for the longitudinal cohort and 246 for the cross-sectional sample unless otherwise specified.
<sup>c</sup>Calculated using the Oxford Multidimensional Poverty Index, which ranges from 0 to 1, with higher scores indicating greater levels of multidimensional poverty. Available at: Oxford Poverty & Human Development Initiative. The Global Multidimensional Poverty Index. https://ophi.org.uk/global-mpi
<sup>d</sup>IPV was administered as part of the routine immunisation schedule at the time when the enrolled 8–9-year-olds were infants (i.e., 2015). The global switch from trivalent OPV to the four-dose bOPV and single-dose IPV schedule was introduced in April 2016.
<sup>e</sup>Rotavac rotavirus vaccine was administered as a three-dose series at 6, 10 and 14 weeks of age.
<sup>f</sup>For the longitudinal cohort, current breastfeeding status was reported by caregivers at enrolment; For the cross-sectional study group, caregivers reported the age at which exclusive breastfeeding was stopped (if stopped) which
was used to calculate breastfeeding status at four months of age, the average age of receipt of the fourth, and final bOPV dose in this cohort.
<sup>g</sup>Amoeba, *Ascaris* worm, Hookworm, Tapeworm, *Hymenolepis nana*.
<sup>h</sup>Enteroinvasive bacterial pathogen associated with dysentery include: Enteroinvasive *Escherichia coli*, Shigella-like toxin-producing *E. coli*, *Campylobacter*, *Salmonella*.
<sup>i</sup>Other bacterial pathogens include: *Clostridium difficile* toxin A/B, *Plesiomonas shigelloides*, Enterotoxigenic *E. coli*, Enteropathogenic *E. coli*, Enteroaggregative *E. coli*.
<sup>j</sup>Viral pathogens include: Rotavirus, Adenovirus, Sapovirus, Astrovirus, Norovirus.
<sup>k</sup>Composite EE score (0–10) calculated from NEO, MPO and AAT concentrations using the method described in Arndt, et al. 2016. [14]. Composite EE scores in the upper quartile ( $\geq 7$ ) were defined as high burden EE.

### Duration of PV1 and PV3 antibodies across longitudinal and cross-sectional study groups

One month post-vaccination, 33% and 18% of participants in the longitudinal and cross-sectional groups had positive stool neutralising activity for PV1, and 66% and 56% positive for PV3, respectively (Figure 2a, Supplementary Table 2). By six months, the proportions were 3% for PV1 in both study groups, and 21% and 27% for PV3, respectively. No participants sampled three or more years after vaccination had positive stool neutralising activity for either serotype. Stool neutralising activity was higher for PV3 than PV1 in both participant groups at all timepoints at which neutralisation was detectable (p<0.00) (Supplementary Table 2). Among infants in the longitudinal cohort, the estimated half-life of stool neutralising activity was 3.4 months (95% confidence interval [CI]: 2.6-5.0) for PV1 and 1.7 months (95% CI: 1.4-2.3) for PV3, derived from samples collected between 1 and 6 months post-vaccination (Supplementary Table 3). Serotype-specific IgA remained detectable for both serotypes among cross-sectionally sampled infants at 3 years and 8-9-years post-vaccination (Figure 2b). Median total stool IgA was similar between enrolment and 6 months post-vaccination for both study groups (range 82.7-112.1 µg/mL) but was <10 µg/mL among cross-sectional study participants vaccinated more than three years previously (Supplementary Table 2).

**Figure 2.**
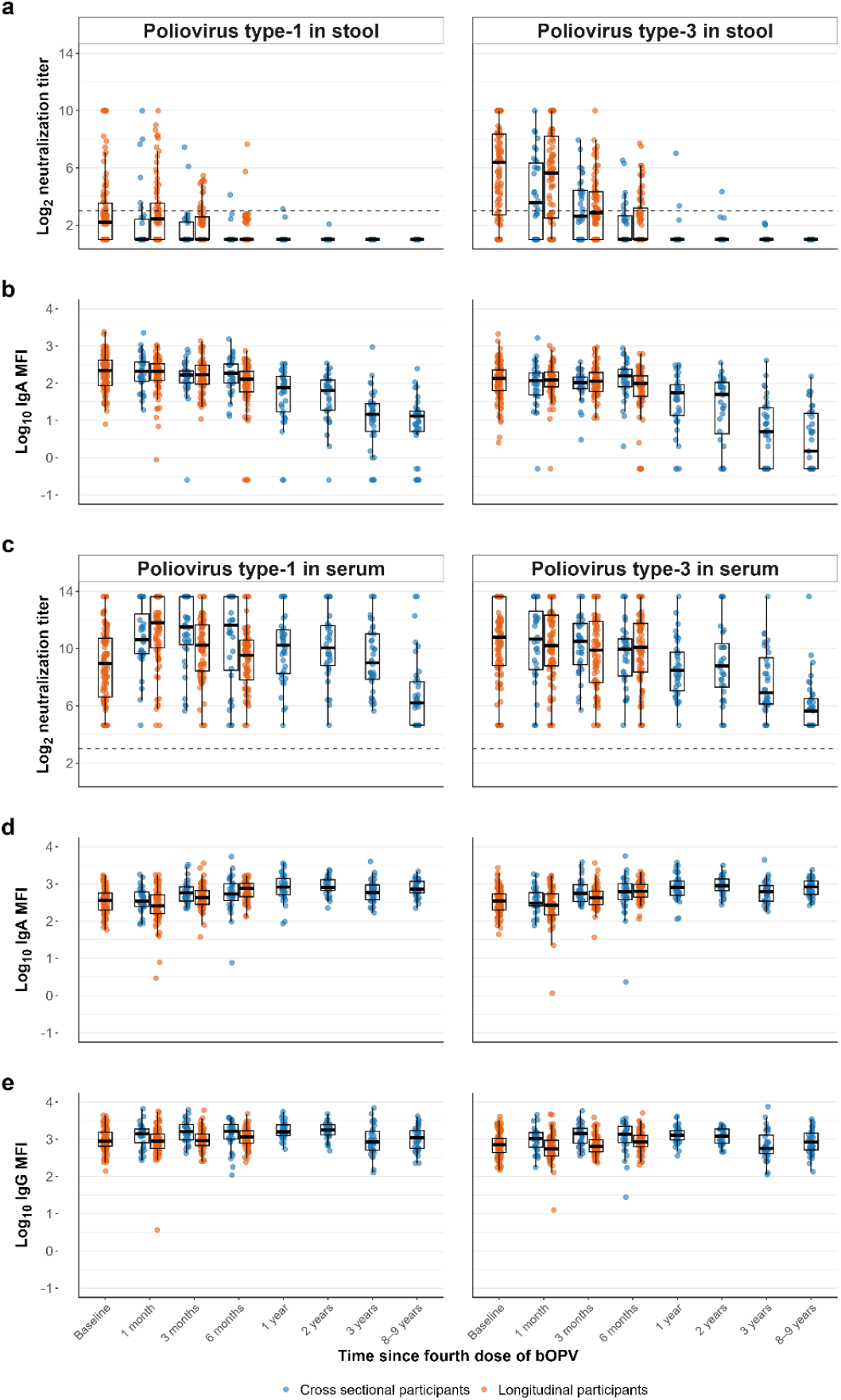
Distribution of poliovirus-specific immune responses following completion of the routine four-dose bOPV vaccination series. Poliovirus type 1 (PV1) and type 3 (PV3)-specific (a) stool log_2_ neutralisation titres (b) log_10_ IgA MFI, (c) serum type-specific log_2_ neutralisation titres, (d) serum log_10_ IgA MFI, and (e) serum log_10_ IgG MFI. IgA and IgG MFI that were negative after background subtraction were assigned a value of half of the lowest observed homotypic MFI. Orange dots represent infants in the longitudinal cohort at baseline and subsequent follow-up visits (n=103), and blue dots represent cross-sectional study participants (n=246). The dashed line represents the positive neutralisation titre threshold (defined as ≥1:8 in the main analysis, equivalent to ≥3 on the log_2_ scale). The time increments on the x-axis are not represented linearly.

All participants were seropositive for both PV1 and PV3 at all timepoints (Figure 2c; Supplementary Table 4). At one month post-vaccination, median serum PV1 log_2_ neutralisation titres were 11.8 in the longitudinal cohort and 10.6 in the cross-sectional group. Corresponding titres for PV3 were 10.2 and 10.7, respectively. Titres remained high from six to 24-months post-vaccination. Serum log_2_ neutralisation titres were lower among children aged 8–9-years the age stratum with the longest time since last vaccination who did not receive IPV (median: 6.2, IQR: 4.6-7.7 for PV1 and 5.6, IQR: 4.6-6.5 for PV3),. Serum neutralising activity for PV1 was equal to or higher than PV3 at all timepoints in both participant groups (Supplementary Table 4). Serum PV-specific IgA levels increased gradually from 1 to 12 months post-vaccination and remained elevated 8-9 years post-vaccination (Figure 2d). Serum PV1 and PV3-specific IgG levels showed little variation across timepoints, including at enrolment, one month, and 8-9 years post-vaccination (Figure 2e).

Modest correlations were observed between PV1 and PV3 log_2_ neutralisation in stool (τ_β_=0.42, p<0.001) and serum (τ_β_=0.31, p<0.001). Homotypic stool and serum neutralisation titres were also modestly correlated (τ_β_=0.32 for both PV1 and PV3, p<0.001) (Supplementary Figure 2). Stool neutralising activity showed modest positive correlations with homotypic stool IgA levels (τ_β_=0.36 for PV1, p<0.001 and τ_β_=0.39 for PV3, p<0.001). All correlations were based on samples collected one month post-vaccination from the longitudinal study group or at enrolment from the cross-sectional study group.

### Kinetics of PV1 and PV3 antibody response among infants in the longitudinal cohort

Among infants in the longitudinal cohort with positive type-specific stool neutralisation at enrolment (29% for PV1 and 72% for PV3), homotypic stool neutralisation declined at each subsequent visit through 6 month follow-up (PV1: 4.8 to 1.0, p<0.001; PV3: 7.3 to 2.2, p<0.001; Figure 3). Among those who were negative for stool neutralisation at enrolment, neutralisation titres increased modestly over the first month (PV1: 1.0 to 2.2; PV3: 1.0 to 2.8) before declining to 1.0 for both serotypes by the 3 month follow-up (p<0.001 for overall change). PV1 serum neutralisation showed an initial increase followed by waning irrespective of PV1 stool neutralisation at enrolment. No strong evidence for a change in PV3-serum neutralisation was observed. Participants with positive PV1 stool neutralisation at enrolment had higher PV1 serum neutralisation titres at enrolment compared to those with negative PV1 stool neutralisation (p=0.0035), and PV1 neutralisation titres were similar at subsequent visits (p>0.2). For PV3, serum neutralisation titres remained higher among those with positive homotypic stool neutralisation at enrolment and through follow-up (p<0.01). Findings were similar in sensitivity analysis restricted to participants who attended all visits (Supplementary Table 5, Supplementary Figure 3). No consistent differences were observed in longitudinal patterns of immunoglobulin levels in stool or serum when comparing positive and negative homotypic stool neutralisation at enrolment.

**Figure 3.**
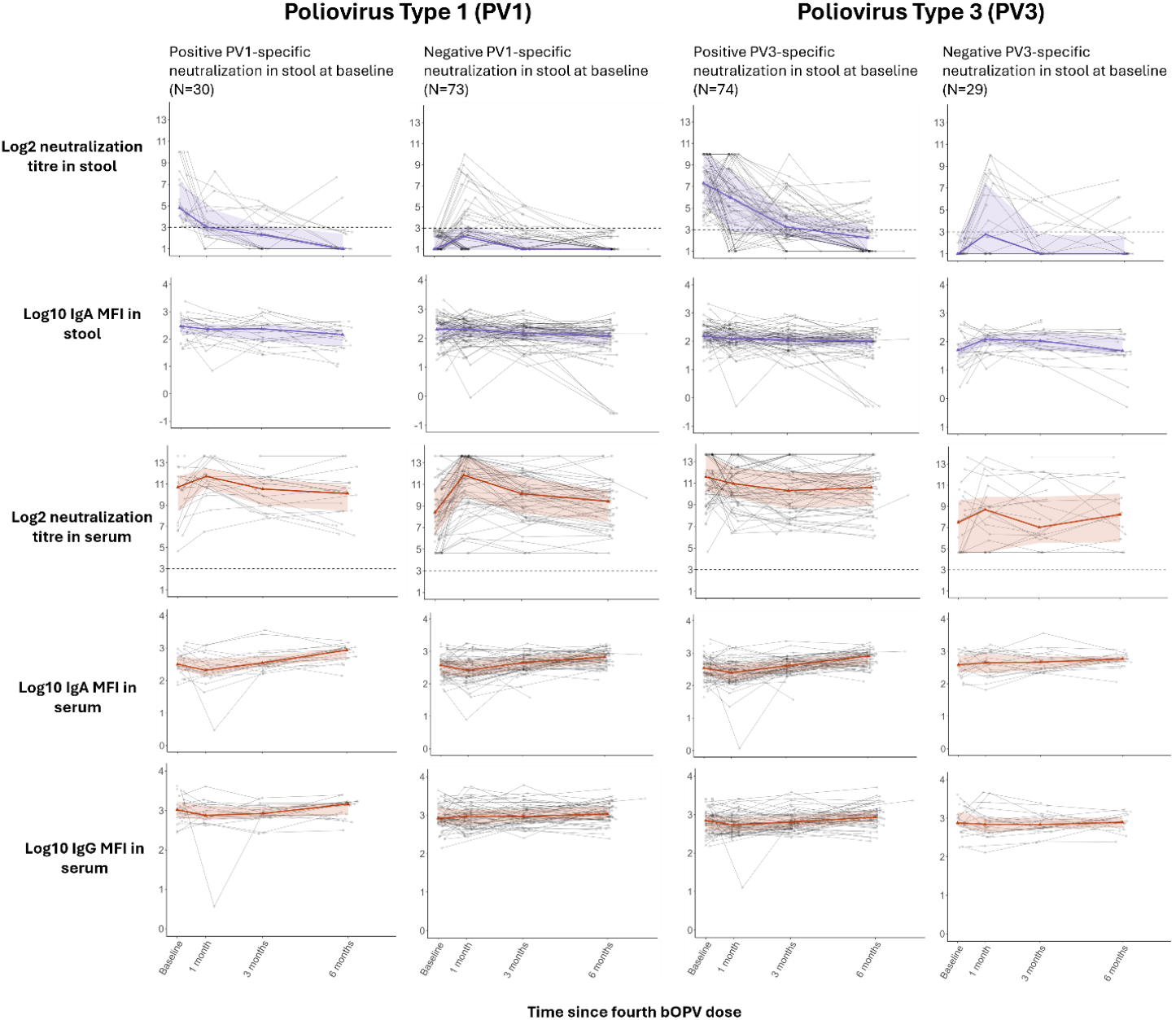
Longitudinal trajectory of type-1 (PV1) and type-3 poliovirus (PV3) specific log_2_ neutralisation titre and log_10_ IgA MFI in stool (purple) and serum (red) samples alongside log_10_ IgG MFI in serum (red) samples among the longitudinal sample (n=103), stratified by whether participants hat positive homotypic neutralisation in stool at baseline (>=1:8). Thirty of the 103 longitudinal participants had positive PV1-specific neutralisation in stool at baseline, and 74 had positive PV3-specific neutralisation in stool at baseline. The bolded line represents the medians at each visit, and the transparent band represents the interquartile range.

### Association between PV1 and PV3 mucosal antibody response, nutritional and coinfection status among infants in the longitudinal cohort

Positive stool neutralisation was detected among 48% of longitudinal cohort participants for PV1 and 84% for PV3 at any visit between enrolment and six month follow-up. There was low evidence for an association between covariates and the odds of positive neutralisation, with the exception of viral coinfection (Figure 4). Concurrent viral infections at the time of polio vaccine series completion tended towards lower odds of positive stool neutralisation for PV1 (adjusted odds ratio (aOR) 0.56, 95% CI 0.25-1.24; p=0.153) and for PV3 (aOR 0.33, 95% CI 0.11-1.01; p=0.052), although confidence intervals included the null. HIV exposure and high EE also tended towards lower odds of positive stool neutralisation for PV3 (aOR 0.29, 95% CI 0.06-1.36; p=0.117 and aOR 0.45, 95% CI 0.15-1.38; p=0.163 respectively), although the estimated effect sizes had low precision and no similar pattern was observed for PV1. Among infants in the longitudinal cohort, PV3 stool neutralisation between 1 and 6 months was observed in 100% of those with schistosome infection at enrolment compared to 82% without (p=0.084). Results were similar when detectable neutralising activity was defied as a titre ≥1:4 rather than ≥1:8, at any timepoint between enrolment and six month follow-up (Supplementary Figure 4).

**Figure 4.**
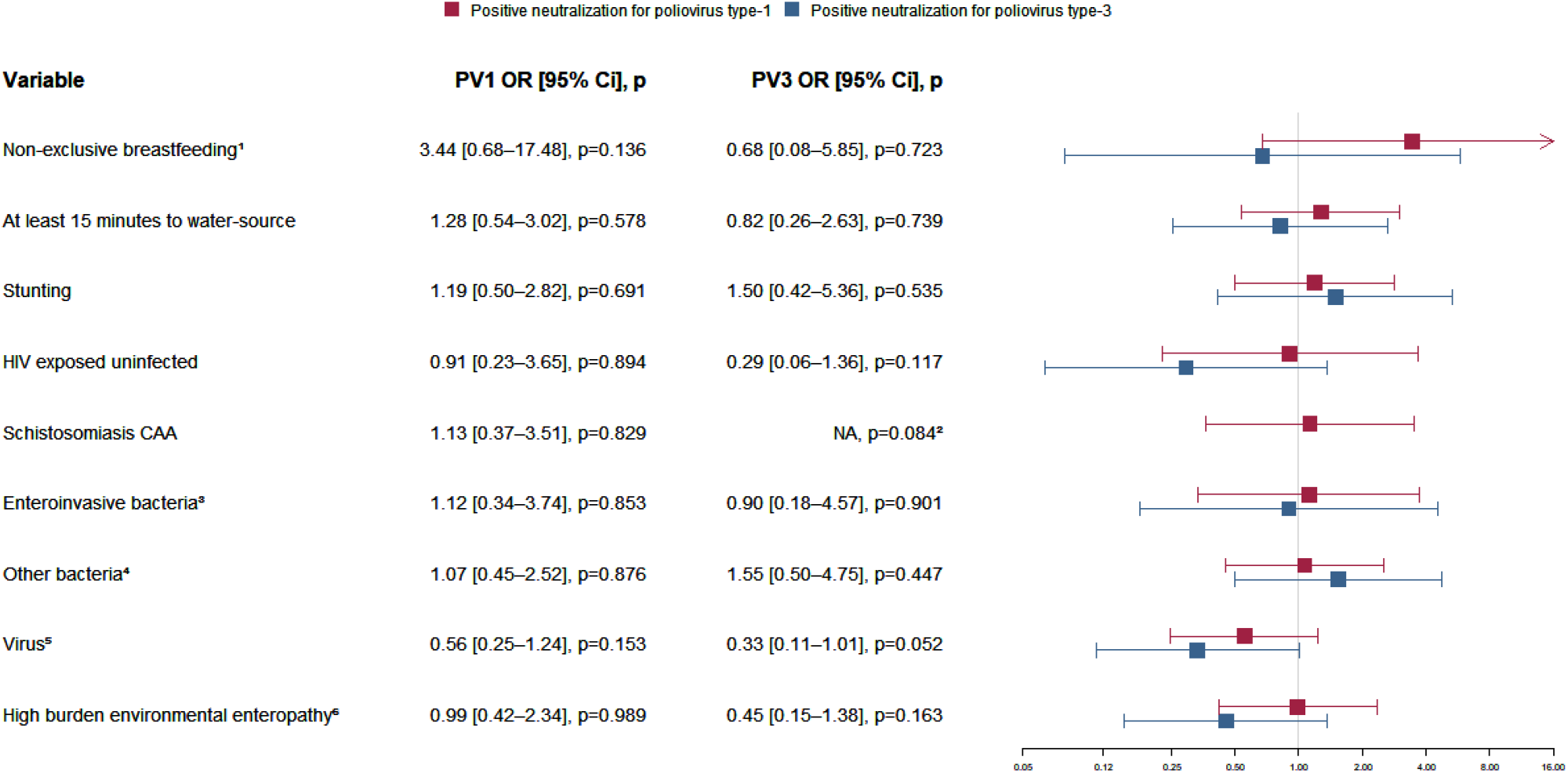
Association between selected characteristics and recording a positive (>=1:8) type-1 (PV1; red) and type-3 poliovirus (PV3; blue) neutralisation titre at any timepoint (between baseline and month-6) among the longitudinal sample (n=103), fitted using a logistic regression adjusted for participant age. The square represents the odds ratio, and the bar represents the 95% confidence interval (CI). ^1^Current breastfeeding status was reported by caregivers at baseline; ^2^All participants who tested positive for schistosome CAA (n=14) also had a positive neutralisation response to type-3 poliovirus. OR not estimable; p-value assessed by Fisher’s exact test; ^3^Includes: Enteroinvasive *E. coli*, Shigella-toxin producing *E. coli, Campylobacter, Salmonella*; ^4^*Clostridium difficile, Plesiomonas shigelloides*, Enterotoxigenic *E. coli*, Enteropathogenic *E. coli*, Enteroaggregative *E. coli;* ^5^Rotavirus, adenovirus, sapovirus, astrovirus, norovirus. ^6^High burden environmental enteropathy (EE) defined as falling in the upper quartile of composite EE scores (i.e. ≥7).

### PV2 neutralisation antibodies among longitudinal and cross-sectional study participants

Positive PV2 stool neutralisation was detected in 6% (22/349) of participants. Stool samples collected at enrolment (from the longitudinal participants) most frequently had positive PV2 neutralisation (13% (13/103) positive), compared to 6% of samples collected from longitudinal and cross-sectional participants 1-month following routine polio immunization completion, and <4% in all later visits (Supplementary Figure 5).

## Discussion

Stool neutralising antibody responses to PV1 and PV3 declined rapidly during the first 3-6 months following completion of the routine four-dose bOPV and single-dose IPV schedule in Tanzania. Among longitudinal cohort participants, the estimated half-life of stool neutralisation was 3.4 months for PV1 and 1.7 months for PV3. Infants in the longitudinal group with positive stool neutralisation at enrolment did not show a boost in homotypic neutralisation following vaccine series completion, and had lower homotypic serum neutralisation responses compared to those with negative neutralisation at enrolment. Despite waning of measurable intestinal antibodies, stool IgA remained detectable 3 years post-vaccination, and serum antibodies declined minimally over the first 2 years.

To our knowledge, this study provides the first direct longitudinal assessment of the duration of mucosal antibody responses after bOPV and IPV beyond 8 weeks. A case series of four infants in Japan reported waning stool IgA, measured using ELISA, 6-34 weeks after two trivalent oral poliovirus vaccine (tOPV) doses, consistent with the waning stool neutralisation observed in this study.^19^ Direct comparisons between IgA patterns is limited by differences in sample dilution and reagent preparation. In addition, previous studies have indicated that the reduction in viral shedding upon subsequent homotypic vaccine challenge declines between ≤1 month and ≥6 months, suggesting waning intestinal immunity.^7, 20^ In contrast, a study in Belgium reported lower shedding after nOPV2 challenge among adults vaccinated with tOPV in infancy compared to unvaccinated adults, indicating long-lasting mucosal immunity, despite no difference in measurable mucosal antibody responses.^6^ It is plausible that unmeasured components of mucosal immunity, such as cell-mediated immunity, may persist into adulthood and contribute to reduced risk of viral infection. Future vaccine challenge studies are needed to understand how trajectories of mucosal antibodies relate to risk of poliovirus infection and viral shedding in the long-term.

Serotype-specific stool neutralisation titres were lower than those reported in previous bOPV trials in Latin America and India,^1, 4, 21^ although were similar to findings in a community-based study among Nigerian children with high bOPV coverage and nOPV2 exposure.^22^ Together, these findings may reflect differences between routine immunisation compared to trial conditions, or known geographical variation in OPV effectiveness.^8^ Most prior studies,^21, 23, 24^ although not all,^19, 22^ have observed stronger PV1 mucosal antibody responses and shedding compared to PV3 after tOPV or bOPV vaccination, possibly reflecting a competitive advantage of PV1 over PV3 during intestinal replication. In contrast, PV3 stool neutralising activity was higher than PV1 in this study. This may result from earlier bOPV doses triggering stronger PV1 intestinal antibody responses initially, allowing PV3 to dominate replication by the time of the fourth bOPV dose. Future longitudinal studies monitoring serotype-specific intestinal antibody responses after successive bOPV doses are needed to clarify how vaccine serotypes compete across immunisation schedules.

Positive PV2 stool neutralisation was detected in 6% of participants, with samples collected at enrolment (from the longitudinal cohort) most frequently positive (Supplementary figure 5). The detection of positive PV2 stool neutralisation may reflect low-level cross-reactivity of bOPV to PV2, or sub-optimal assay specificity. Alternatively, some participants may have had asymptomatic infection with cVDPV2, which was detected in Tanzania during the study period through environmental surveillance.^25^ It is possible that some older participants may have been exposed to the subnational 2023 nOPV2 campaign, although the study region does not neighbour any campaign regions and it was not possible to verify nOPV2 exposure status which is not recorded on vaccination cards.

Longitudinal cohort participants with positive stool neutralisation at enrolment showed no evidence of homotypic boosting and had minimal changes in serum neutralisation. Participants who were negative at enrolment showed weak increases in stool and serum neutralisation one-month post-vaccination. Negative stool neutralisation at enrolment might reflect limited responses to the prior three bOPV doses or could indicate waning of short-lived intestinal antibody responses. No boost in stool IgA was observed except for an increase in PV3-specific IgA among participants who were negative for PV3 stool neutralisation at baseline. Serum IgA and IgG increased gradually between 1 and 6 months post-vaccination, suggesting a slower serum response to OPV and IPV. Previous studies have similarly found limited intestinal antibody boosting upon successive OPV doses with short vaccine intervals, particularly among children with existing mucosal antibody responses.^2, 4^ This evidence suggests that while successive OPV doses may augment intestinal mucosal immunity among those who previously missed vaccination or experienced low vaccine ‘take’ (intestinal replication), OPV may not play a major role in boosting intestinal antibody titres among prior responders.

The high prevalence of gastrointestinal enteric infections and high burden of EE at the time of bOPV vaccination, consistent with nationwide evidence,^26, 27^ highlights the complex intestinal environment in which immune responses to oral vaccines develop in pathogen-endemic settings. Concomitant enteric infections and EE have been hypothesised to reduce oral vaccine performance through several mechanisms, including disruption of gut barrier integrity, microbial competition at viral binding sites, and heightened non-specific immune activation that may limit vaccine virus replication.^8, 28^ To our knowledge, this is the first study to directly evaluate associations between gastrointestinal coinfections and vaccine-induced intestinal antibody responses in a community setting. Previous studies have demonstrated associations between non-polio enterovirus infections and reduced OPV seroconversion or increased viral shedding, though evidence remains inconsistent.^8, 29^ Our results indicate that the negative association between enterovirus co-infections and OPV responses extends to intestinal antibody responses. We observed no clear associations with non-exclusive breastfeeding, stunting, or other co-infections, although power to detect differences was limited. High burden EE and HIV exposure tended towards weaker PV3, but not PV1, responses with low statistical evidence for the associations. This suggests a potential serotype-specific effect of co-infections on OPV responses, consistent with previous studies.^29, 30^

Our study has several limitations. First, the first scheduled follow-up was one-month post-vaccination, which may underrepresent antibody responses that peaked earlier; previous trials have found robust mucosal antibody responses to OPVs within two weeks.^4, 6^ Second, the neutralisation assay has limited sensitivity and may not detect low-level neutralisation, which may could still confer functional immunity and protection from future infection. Third, variations in assay performance mean that neutralisation titres, and immunoglobulin MFI cannot be directly compared between studies. Additionally, antibodies detected in stool may have derived from maternal breastmilk rather than from the infants’ vaccine response; however, we observed no association between exclusive breastfeeding and neutralising antibodies. Coinfection and nutritional status were assessed only at the time of fourth bOPV dose receipt, with no measurements available for earlier doses. Finally, statistical power to detect associations between mucosal responses and covariates was limited, meaning that small but potentially meaningful associations may have been missed.

Intestinal PV specific neutralisation was low and waned within 6 months after completion of the routine four-dose bOPV and single-dose IPV schedule among children in Tanzania. Detectable serum neutralisation had a longer duration. Mucosal antibody responses are likely important in rapid functional immunity immediately after vaccination. Previous evidence indicates that some level of immune protection against infection persists into adulthood, although the underlying immunological mechanism remains unclear. Future research is needed to determine optimal vaccine strategies to achieve durable mucosal immunity, and to evaluate how the duration of vaccine-induced intestinal antibodies is correlated with protection against future poliovirus infection.

## Acknowledgements

The authors wish to thank all the children that participated in this study and their parent(s)/guardians for their time and trust. The authors gratefully acknowledge the study nurses that made this work possible: Ndalloh Paul, Teddy Zephinia, Suzan Pungu, Maneno Siwga. The authors also appreciate the work of the parasitologists, Josephat Kuboza, and Petero Mnyeshi.

## Authors’ contributions

AAC – project administration, investigation, data curation, formal analysis, visualisation, writing – original draft; NM – formal analysis, visualisation, writing – original draft; MHL – formal analysis, supervision, validation, writing – review and editing; MEA resources, software, writing – review and editing; JAW – software, investigation, writing – review and editing; SS – investigation, writing – review and editing; OM – project administration, investigation, writing – review and editing; EM - project administration, investigation, writing – review and editing; LM – investigation, writing – review and editing; NU investigation, writing – review and editing; AG – formal analysis, writing – review and editing; MA – methodology, writing – review and editing; RMB – conceptualisation, writing – review and editing; OBLC – investigation, writing – review and editing; DDJ –investigation, writing – review and editing; PTH – investigation, validation, writing – review and editing; GJVD – resources, investigation, validation, writing – review and editing; PC – resources, investigation, validation, writing – review and editing; MW – investigation, writing – review and editing; LFW resources, investigation, supervision, writing – review and editing; AH – supervision, writing – review and editing; SK – supervision, writing – review and editing; WWA – investigation, writing review and editing; RIC – investigation, writing – review and editing; EBB – methodology, supervision, writing – original draft; JAD – conceptualisation, methodology, validation, supervision, writing - original draft; PFW - conceptualisation, methodology, supervision, funding acquisition, writing - original draft.

All authors had full access to all data in the study, read the drafted manuscript, provided feedback, and had final responsibility for the decision to submit for publication. AAC, NM, MHL, EBB, JAD, and PFW have all directly accessed and verified the underlying data reported in the manuscript.

## Data availability

De-identified data can be made available to qualified investigators upon reasonable request to the corresponding author. The analysis code is publicly available at https://github.com/Naomi-miall/Duration-of-mucosal-immunity-following-routine-bOPV-immunisation-in-Tanzania.

## References

1. Connor, R.I., et al., Mucosal immunity to poliovirus. Mucosal Immunology, 2022. 15(1): p. 1–9.

2. John, J., et al., Effect of a single inactivated poliovirus vaccine dose on intestinal immunity against poliovirus in children previously given oral vaccine: an open-label, randomised controlled trial. The Lancet, 2014. 384(9953): p. 1505–1512.

3. Gamage, D., et al., Achieving high seroprevalence against polioviruses in Sri Lanka--results from a serological survey, 2014. J Epidemiol Glob Health, 2015. 5(4 Suppl 1): p. S67–71.

4. Brickley, E.B., et al., Intestinal Immunity to Poliovirus Following Sequential Trivalent Inactivated Polio Vaccine/Bivalent Oral Polio Vaccine and Trivalent Inactivated Polio Vaccine-only Immunization Schedules: Analysis of an Open-label, Randomized, Controlled Trial in Chilean Infants. Clin Infect Dis, 2018. 67(suppl_1): p. S42–s50.

5. Wright, P.F., et al., Intestinal Immunity Is a Determinant of Clearance of Poliovirus After Oral Vaccination. The Journal of Infectious Diseases, 2014. 209(10): p. 1628–1634.

6. Godin, A., et al., Intestinal mucosal immune responses induced by novel oral poliovirus vaccine type 2 and Sabin monovalent oral poliovirus vaccine type 2: an analysis of data from four clinical trials. Lancet Microbe, 2025: p. 101028.

7. Grassly, N.C., et al., Waning Intestinal Immunity After Vaccination With Oral Poliovirus Vaccines in India. The Journal of Infectious Diseases, 2012. 205(10): p. 1554–1561.

8. Parker, E.P.K., et al., Causes of Impaired Oral Vaccine Efficacy in Developing Countries. Future Microbiology, 2018. 13(1): p. 97–118.

9. Patriarca, P.A., P.F. Wright, and T.J. John, Factors Affecting the Immunogenicity of Oral Poliovirus Vaccine in Developing Countries: Review. Reviews of Infectious Diseases, 1991. 13(5): p. 926–939.

10. Mnkugwe, R.H., et al., Prevalence and correlates of intestinal schistosomiasis infection among school-aged children in North-Western Tanzania. PLOS ONE, 2020. 15(2): p. e0228770.

11. Bosch, F., et al., Diagnosis of soil-transmitted helminths using the Kato-Katz technique: What is the influence of stirring, storage time and storage temperature on stool sample egg counts? PLoS Negl Trop Dis, 2021. 15(1): p. e0009032.

12. Corstjens, P.L.A.M., et al., Tools for diagnosis, monitoring and screening of Schistosoma infections utilizing lateral-flow based assays and upconverting phosphor labels. Parasitology, 2014. 141(14): p. 1841–1855.

13. Arndt, M.B., et al., Fecal Markers of Environmental Enteropathy and Subsequent Growth in Bangladeshi Children. Am J Trop Med Hyg, 2016. 95(3): p. 694–701.

14. Kosek, M.N., Causal Pathways from Enteropathogens to Environmental Enteropathy: Findings from the MAL-ED Birth Cohort Study. EBioMedicine, 2017. 18: p. 109–117.

15. Buss, S.N., et al., Multicenter evaluation of the BioFire FilmArray gastrointestinal panel for etiologic diagnosis of infectious gastroenteritis. J Clin Microbiol, 2015. 53(3): p. 915–25.

16. Arita, M., et al., Development of a poliovirus neutralization test with poliovirus pseudovirus for measurement of neutralizing antibody titer in human serum. Clin Vaccine Immunol, 2011. 18(11): p. 1889–94.

17. Word Health Organization, WHO Anthro Survey Analyser: Software for analysing survey anthropometric data for children under 5 years of age. Built-in software edition. Version 1.0. 2018: Geneva.

18. Obuchowski, N.A., On the comparison of correlated proportions for clustered data. Stat Med, 1998. 17(13): p. 1495–507.

19. Nishio, O., et al., Fecal IgA Antibody Responses after Oral Poliovirus Vaccination in Infants and Elder Children. Microbiology and Immunology, 1990. 34(8): p. 683–689.

20. John, J., et al., The Duration of Intestinal Immunity After an Inactivated Poliovirus Vaccine Booster Dose in Children Immunized With Oral Vaccine: A Randomized Controlled Trial. J Infect Dis, 2017. 215(4): p. 529–536.

21. Sutter, R.W., et al., Immunogenicity of bivalent types 1 and 3 oral poliovirus vaccine: a randomised, double-blind, controlled trial. Lancet, 2010. 376(9753): p. 1682–8.

22. Cooper, L., et al., Mucosal Immune Responses to Novel Oral Poliovirus Vaccine Type-2 (nOPV2) Supplementary Immunization Activities in Northern Nigeria: A Community-Based Cohort Study [Pre-print].

23. Keller, R., et al., Intestinal IgA neutralizing antibodies in newborn infants following poliovirus immunization. Pediatrics, 1969. 43(3): p. 330–8.

24. Bhaskaram, P., et al., Systemic and mucosal immune response to polio vaccination with additional dose in newborn period. J Trop Pediatr, 1997. 43(4): p. 232–4.

25. Global Polio Eradication Initiative (GPEI). Circulating Vaccine-Derived Poliovirus Count. 2026 [cited 2026 29/01/2026]; Available from: https://polioeradication.org/circulating-vaccine-derived-poliovirus-count/.

26. Piovani, D., et al., The global burden of enteric fever, 2017–2021: a systematic analysis from the global burden of disease study 2021. eClinicalMedicine, 2024. 77.

27. Cordeiro, A., et al., Intestinal Parasitic Infections in Early Infancy: A Longitudinal Report of Schistosoma and Entamoeba Infections Among Infants in Tanzania [Version 1]. VeriXiv, 2026. 3: p. 77.

28. The MAL-ED Network Investigators, The MAL-ED Study: A Multinational and Multidisciplinary Approach to Understand the Relationship Between Enteric Pathogens, Malnutrition, Gut Physiology, Physical Growth, Cognitive Development, and Immune Responses in Infants and Children Up to 2 Years of Age in Resource-Poor Environments. Clinical Infectious Diseases, 2014. 59(suppl_4): p. S193–S206.

29. Taniuchi, M., et al., Impact of enterovirus and other enteric pathogens on oral polio and rotavirus vaccine performance in Bangladeshi infants. Vaccine, 2016. 34(27): p. 3068–3075.

30. Parker, E.P.K., et al., Influence of Enteric Infections on Response to Oral Poliovirus Vaccine: A Systematic Review and Meta-analysis. The Journal of Infectious Diseases, 2014. 210(6): p. 853–864.

