## Supplementary Tables and Figures for "Duration of intestinal mucosal antibody responses to poliovirus in children routinely immunised with bivalent oral polio vaccine and inactivated polio vaccine in Tanzania: A longitudinal cohort and cross-sectional study"

**Supplementary materials:**

| **Supplementary Table 1:** Enteric pathogens detected by multiplex PCR assay (BIOFIRE FILMARRAY Gastrointestinal panel) at baseline (<7 days after fourth and final bOPV vaccine) among infants in the longitudinal cohort (N=103). | |
|---|---|
| *Variable* | % (n) |
| **Bacteria** | |
| *Campylobacter* (*C. coli*, *C. jejuni*, *C. upsaliensis*) | 5.8% (6) |
| *Clostridium difficile* toxin A/B | 2.9% (3) |
| *Plesiomonas shigelloides* | 2.9% (3) |
| *Salmonella* | 5.8% (6) |
| *Vibrio* (*V. cholerae*, *V. parahaemolyticus*, *V. vulnificus*) | 0 (0%) |
| *Yersinia enterocolitica* | 0 (0%) |
| **Diarrheagenic *E. coli/Shigella*** | |
| Enteroaggregative *Escherichia coli* | 67.0% (69) |
| Enteropathogenic *E. coli* | 19.4% (20) |
| Enterotoxigenic *E. coli lt/st* | 10.7% (11) |
| *Shigella*-like toxin-producing *E. coli stx1/stx2* (including *Escherichia coli* O157) | 1.9% (2) |
| *Shigella*/Enteroinvasive *E. coli* | 0.0% (0) |
| **Parasites** | |
| *Cryptosporidium* | 0 (0%) |
| Cyclospora *cayetanensis* | 0 (0%) |
| *Entamoeba histolytica* | 0 (0%) |
| *Giardia lamblia* | 4.9% (5) |
| **Viruses** | |
| Adenovirus F 40/41 | 1.9% (2) |
| Astrovirus | 2.9% (3) |
| Norovirus GI/GII | 12.6% (13) |
| Rotavirus A | 26.2% (27) |
| Sapovirus | 2.9% (2) |
| No pathogen detected | 23 (22.3%) |
| One pathogen detected | 31 (30.1%) |
| Two pathogens detected | 22 (21.4%) |
| Three or more pathogens detected | 27 (26.2%) |

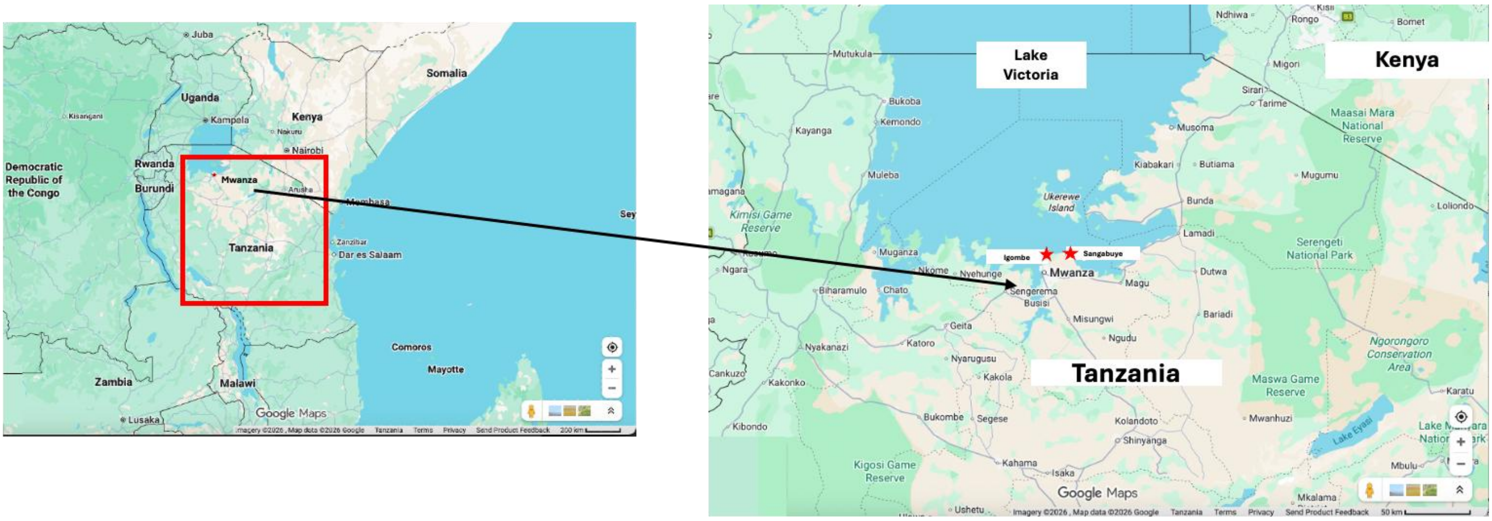

**Supplementary Figure 1:** Map of Mwanza, Tanzania showing the two lakeside study communities that children were recruited from, Igombe and Sangabuye.

**Supplementary Table 2:** Poliovirus type-1 and type-3 (PV1 and PV3) specific stool neutralising activity and stool IgA MFI, and total IgA measured at the time of, and 1-month to 8-9-years following, the fourth bOPV dose and first IPV dose in the routine immunisation schedule among a longitudinal cohort and a cross-sectional sample of infants and children in Tanzania. P-values for overall change across timepoints within each cohort and poliovirus serotype are shown in the bottom row for each model and were calculating using a Wald test after fitting a mixed-effect logistic regression (for positive stool neutralising activity) or mixed-effect linear regression (for neutralising activity titre or MFI), modelling visit as a factor variable and individual participant as a random-effect to account for repeated observations in the longitudinal cohort. Within-individual comparisons between PV1 and PV3 positive neutralisation (binary outcomes) were analysed using an extension of McNemar's test to account for repeated observations (right column).[18] Differences in PV1 and PV3 log-transformed neutralisation titres(continuous outcome), were modelled using mixed-effects linear regression with participant-level random effects.

| | | **Longitudinal (N=103)** | | **Cross-sectional (N=246)** | | **p-value comparing PV1 and PV3** |
|---|---|---|---|---|---|---|
| | | **PV1** | **PV3** | **PV1** | **PV3** | |
| **Positive stool neutralising activity (>=1:8) % (n/N)** | Baseline | 29% (30/103) | 72% (74/103) | Not collected | | <0.001 |
| | 1-month | 33% (27/82) | 66% (54/82) | 18% (6/34) | 56% (19/34) | |
| | 3-months | 18% (14/77) | 47% (36/77) | 6% (2/35) | 46% (16/35) | |
| | 6-months | 3% (2/77) | 27% (21/77) | 3% (1/33) | 21% (7/33) | |
| | 12-months | Not collected | | 3% (1/38) | 5% (2/38) | |
| | 24-months | | | 0% (0/32) | 3% (1/32) | |
| | 36-months | | | 0% (0/37) | 0% (0/37) | |
| | 8-9-years | | | 0% (0/37) | 0% (0/37) | |
| | p-value | <0.001 | <0.001 | 0.098 | <0.001 | |
| **Stool neutralising activity (log$_2$ transformed) Median (IQR)** | Baseline | 2.2 (1.0-3.6), n=103 | 6.4 (2.6–8.4), n=103 | Not collected | | <0.001 |
| | 1-month | 2.4 (1.0-3.6), n=82 | 5.6 (2.5 – 8.3), n=82 | 1.0 (1.0-2.5), n=34 | 3.6 (1.0-6.5), n=34 | |
| | 3-months | 1.0 (1.0-2.6), n=77 | 2.8 (1.0 – 4.3), n=77 | 1.0 (1.0-2.3), n=35 | 2.6 (1.0-4.5), n=35 | |
| | 6-months | 1.0 (1.0 – 1.0), n=77 | 1.0 (1.0 – 3.2), n=77 | 1.0 (1.0-1.0), n=33 | 1.0 (1.0-2.6), n=33 | |
| | 12-months | Not collected | | 1.0 (1.0-1.0), n=38 | 1.0 (1.0-1.0), n=38 | |
| | 24-months | | | 1.0 (1.0-1.0), n=32 | 1.0 (1.0-1.0), n=32 | |
| | 36-months | | | 1.0 (1.0-1.0), n=37 | 1.0 (1.0-1.0), n=37 | |
| | 8-9-years | | | 1.0 (1.0-1.0), n=37 | 1.0 (1.0-1.0), n=37 | |
| | p-value | <0.001 | <0.001 | <0.001 | <0.001 | |

| | | | | | | |
|---|---|---|---|---|---|---|
| **Stool IgA MFI ($log_{10}$ transformed) Median (IQR)** | Baseline | 2.3 (1.9-2.6), n=102 | 2.1 (1.8-2.4), n=102 | Not collected | | NA |
| | 1-month | 2.3 (2.1-2.5), n=82 | 2.1 (1.9-2.3), n=82 | 2.3 (2.0-2.6) n=34 | 2.1 (1.7-2.3) n=34 | |
| | 3-months | 2.2 (2.0-2.5), n=77 | 2.1 (1.8-2.3), n=77 | 2.2 (2.0-2.3) n=35 | 2.0 (1.8-2.2) n=35 | |
| | 6-months | 2.1 (1.8-2.3), n=77 | 2.0 (1.6-2.2), n=77 | 2.3 (2.0-2.5) n=33 | 2.2 (1.9-2.4) n=33 | |
| | 12-months | Not collected | | 1.9 (1.2-2.2) n=37 | 1.7 (1.1-2.0) n=37 | |
| | 24-months | | | 1.8 (1.3-2.1) n=32 | 1.7 (0.6-2.0) n=32 | |
| | 36-months | | | 1.2 (0.7-1.4) n=37 | 0.7 (-0.3-1.3) n=37 | |
| | 8-9-years | | | 1.1 (0.7-1.3) n=37 | 0.2 (-0.3-1.2) n=37 | |
| | p-value | <0.001 | <0.001 | <0.001 | <0.001 | |
| **Stool Total IgA µg/mL Median (IQR)** | Baseline | 82.7 (54.4-157.2), n=99 | | Not collected | | NA |
| | 1-month | 111.8 (59.1-173.7), n=78 | | 97.0 (43.6-143.3) n=33 | | |
| | 3-months | 98.3 (68.1-166.6), n=77 | | 112.1 (65.3-144.5) n=34 | | |
| | 6-months | 86.2 (47.3-161.2), n=73 | | 108.5 (68.0-224.4) n=32 | | |
| | 12-months | Not collected | | 51.5 (23.2-84.5) n=34 | | |
| | 24-months | | | 33.9 (21.4-52.0) n=25 | | |
| | 36-months | | | 8.0 (6.0-28.1) n=21 | | |
| | 8-9-years | | | 4.3 (3.4-5.3) n=17 | | |
| | p-value | 0.7007 | | 0.0149 | | |

PV1 – Poliovirus Type-1; PV3 – Poliovirus Type-3; IQR – Interquartile Range; MFI – Median Fluorescence Intensity

**Supplementary Table 3**: Estimated linear regression co-efficient, associated p-value and half-life for poliovirus type-1 and type-3 (PV1 and PV3) $log_2$-neutralisation titre among the longitudinal cohort calculated using a mixed-effects linear regression model fitting stool $log_2$-neutralisation titres from stool and serum samples collected 1- , 3- and 6-months post vaccination, adjusted for age as a fixed-effect and individual participant as a random-effect.

| | **Linear regression co-efficient (95% CI)** | **P-Value** | **Half-life in months (95% CI)** |
|---|---|---|---|
| **PV1 in stool** | | | |
| Longitudinal cohort[a] | -0.3 (-0.4 to -0.2) | <0.001 | 3.4 (2.6 to 5.0) |
| Participants who were positive (≥1:8) for PV1 stool neutralisation at baseline | -0.3 (-0.5 to -0.1) | 0.002 | 3.0 (1.8 to 8.1) |
| Participants who were negative (<1:8) for PV1 stool neutralisation at baseline | -0.3 (-0.4 to -0.2) | <0.001 | 3.6 (2.6 to 5.6) |
| **PV3 in stool** | | | |
| Longitudinal cohort[a] | -0.6 (-0.7 to -0.4) | <0.001 | 1.7 (1.4 to 2.3) |
| Participants who were positive (≥1:8) for PV3 stool neutralisation at baseline | -0.7 (-0.8 to -0.5) | <0.001 | 1.5 (1.2 to 2.0) |
| Participants who were negative (<1:8) for PV3 stool neutralisation at baseline | -0.4 (-0.7 to -0.3) | 0.004 | 2.5 (1.5 to 8.0) |

[a]Includes those with missing follow-up study visits.

PV1 – Poliovirus Type-1; PV3 – Poliovirus Type-3; CI – Confidence Interval

**Supplementary Table 4:** Poliovirus type-1 and type-3 (PV1 and PV3) specific serum neutralising activity and serum IgA and IgG MFI measured at the time of, and 1-month to 8-9 years following completion of the routine polio vaccine immunisation schedule among a longitudinal and a cross-sectional sample of infants and children in Tanzania. P-values for overall change across timepoints within each cohort and poliovirus serotype, shown in the bottom row for each model, were calculated using a Wald test after fitting a mixed-effect linear regression, modelling visit as a factor variable and individual participant as a random-effect to account for repeated observations in the longitudinal cohort. Statistical evidence for differences in PV1 and PV3 log2-transformed neutralisationwas assessed using mixed-effects linear regression models with participant-level random effects.

| | | **Longitudinal cohort (N=103)** | | **Cross-sectional study participants (N=246)** | |
|---|---|---|---|---|---|
| | | **PV1** | **PV3** | **PV1** | **PV3** |
| **Serum neutralising activity ($log_2$ transformed) Median (IQR)** | Baseline | 9.0 (6.6-10.7),n=97 | 10.8 (8.8-13.6), n=97 | Not collected | |
| | 1-month | 11.8 (10.0-13.6),n=78 | 10.2 (8.8-12.4), n=78 | 10.6 (9.6-12.8), n=32 | 10.7 (8.5-13.0), n=32 |
| | 3-months | 10.2 (8.4-11.7),n=75 | 9.9 (7.6-11.9), n=75 | 11.5 (10.1-13.6),n=35 | 10.5 (8.8-11.8), n=35 |
| | 6-months | 9.5 (7.6-10.6),n=71 | 10.1 (8.3-11.8), n=71 | 11.7 (8.4-13.6), n=32 | 10.0 (8.0-10.7), n=32 |
| | 12-months | Not collected | | 10.2 (8.3-11.3), n=37 | 8.5 (7.1-9.7), n=37 |
| | 24-months | | | 10.1 (8.8-11.7), n=32 | 8.8 (7.2-10.6), n=32 |
| | 36-months | | | 9.0 (7.8-11.1), n=36 | 6.9 (6.1-9.5), n=36 |
| | 8-9-years | | | 6.2 (4.6-7.7), n=37 | 5.6 (4.6-6.5), n=37 |
| | p-value | <0.001 | 0.045 | <0.001 | <0.001 |
| p-value comparing PV1 and PV3 serum neutralising activity | | 0.001 | | | |
| **Serum IgA MFI ($log_{10}$ transformed) Median (IQR)** | Baseline | 2.6 (2.3-2.8), n=94 | 2.5 (2.3-2.7), n=94 | Not collected | |
| | 1-month | 2.4 (2.2-2.7), n=78 | 2.4 (2.2-2.7), n=78 | 2.5 (2.4-2.8) n=34 | 2.5 (2.4-2.8) n=34 |
| | 3-months | 2.6 (2.4-2.8), n=75 | 2.6 (2.4-2.8), n=75 | 2.8 (2.5-2.9) n=35 | 2.7 (2.5-3.0) n=35 |
| | 6-months | 2.9 (2.7-3.0), n=71 | 2.8 (2.6-3.0), n=71 | 2.7 (2.5-3.0) n=33 | 2.8 (2.6-3.0) n=33 |
| | 12-months | Not collected | | 2.9 (2.7-3.2) n=37 | 2.9 (2.7-3.1) n=37 |
| | 24-months | | | 2.9 (2.8-3.1) n=32 | 3.0 (2.8-3.1) n=32 |
| | 36-months | | | 2.8 (2.6-3.0) n=37 | 2.8 (2.5-3.0) n=37 |
| | 8-9-years | | | 2.9 (2.8-3.1) n=37 | 2.9 (2.7-3.1) n=37 |
| | p-value | <0.001 | <0.001 | <0.001 | <0.001 |

| | | | | | |
|---|---|---|---|---|---|
| **Serum IgG MFI (log$_{10}$ transformed) Median (IQR)** | Baseline | 2.9 (2.8-3.2), n=95 | 2.9 (2.6-3.0), n=95 | Not collected | |
| | 1-month | 2.9 (2.7-3.1), n=78 | 2.7 (2.5-3.0), n=78 | 3.1 (2.9-3.3) n=34 | 3.0 (2.8-3.2) n=34 |
| | 3-months | 3.0 (2.8-3.1), n=75 | 2.8 (2.7-3.0), n=75 | 3.2 (3.0-3.4) n=35 | 3.2 (2.9-3.3) n=35 |
| | 6-months | 3.1 (2.9-3.2), n=72 | 2.9 (2.8-3.1), n=72 | 3.2 (3.0-3.5) n=33 | 3.1 (2.9-3.4) n=33 |
| | 12-months | Not collected | | 3.2 (3.1-3.4) n=37 | 3.1 (3.0-3.2) n=37 |
| | 24-months | | | 3.3 (3.1-3.4) n=32 | 3.1 (2.9-3.3) n=32 |
| | 36-months | | | 2.9 (2.7-3.2) n=37 | 2.8 (2.6-3.1) n=37 |
| | 8-9-years | | | 3.0 (2.8-3.2) n=37 | 2.9 (2.7-3.2) n=37 |
| | p-value | 0.081 | <0.001 | <0.001 | 0.001 |

PV1 – Poliovirus Type-1; PV3 – Poliovirus Type-3; IQR – Interquartile Range; MFI – Median Fluorescence Intensity

**Supplementary Figure 2**:

Pairwise correlation between a) poliovirus types-1 and -3 specific $log_2$ neutralising titres in stool and serum samples; b) homotypic stool and serum $log_2$ neutralising titres; c) homotypic $log_2$ neutralising titres and $log_{10}$ IgA MFI in stool; d) $log_{10}$ IgA MFI serotype-specific IgA MFI and total IgA ug/ml in stool; e) homotypic $log_2$ neutralising titres and $log_{10}$ IgA MFI in serum; f) homotypic $log_2$ neutralising titres and $log_{10}$ IgG MFI in serum; g) homotypic $log_{10}$ IgA MFI and $log_{10}$ IgG MFI in serum. For panels b) to g) the left-hand column shows poliovirus type-1 specific data and the right-hand column shows poliovirus type-3 specific data. Blue points represent cross-sectional sample participants (n=246) and orange points represent longitudinal sample participants when measured at 1-month after fourth bOPV dose (n=84). Correlation coefficients are estimated from Kendall's tau-b correlations.

Stool samples Serum samples

a) PV1- and PV3-specific log2 neutralization titer

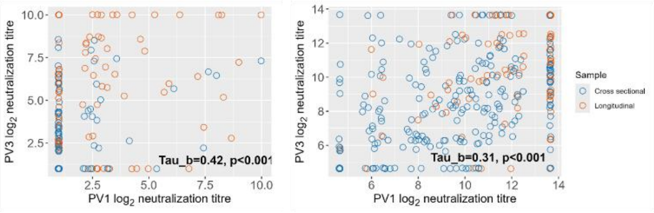

PV1-specific PV3-specific

b) Log2 neutralization titer in stool and serum

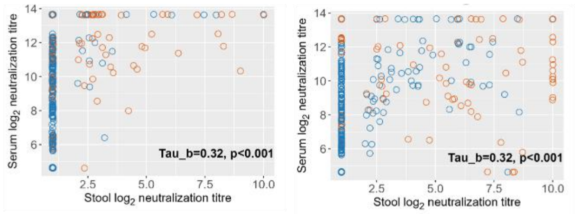

### Stool

c) Log2 neutralization titer and log10 serotype-specific IgA MFI

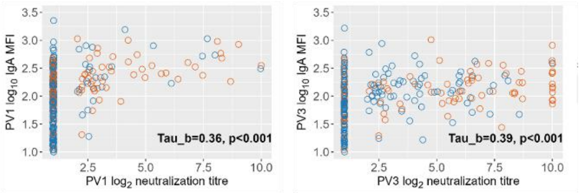

d) Log10 serotype-specific IgA MFI and total IgA ug/ml

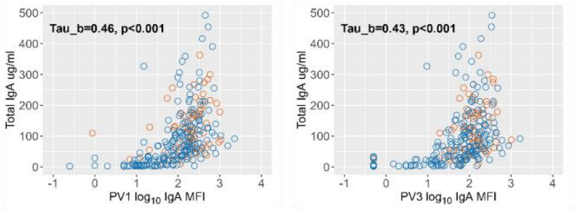

### Serum

e) Log2 neutralization titer and log10 serotype-specific IgA MFI

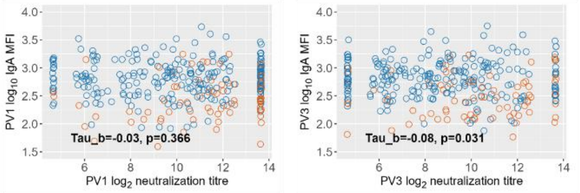

f) Log2 neutralization titer and log10 serotype-specific IgG MFI

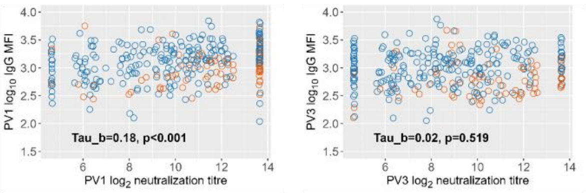

g) Log10 serotype-specific IgA MFI and log10 serotype-specific IgG MFI

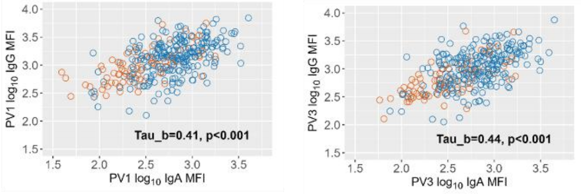

**Supplementary Table 5:** Sociodemographic characteristics of participants with complete data in the longitudinal cohort and the full cohort including those with missing data:

| Variable (at baseline interview) | Full longitudinal cohort N=103[a] | Longitudinal cohort with complete data N=66[a] |
|---|---|---|
| Female | 49.5% (51) | 51.5% (34) |
| Age of mother at participant birth (years) | 26 [21-33] | 28 [22-32] |
| Any adult in household completed ≥ 6 years of education | 43.7% (45) | 37.9% (25) |
| Distance to water source (minutes walking) | 5 [3-15] | 9 [5-15] |
| Multidimensional Poverty Index Score[b] (ranges from 0-1, with 1 indicating greater multidimensional poverty) | 0.72 [0.61-0.83], n=94 | |
| **Vaccine history** | | |
| Age (weeks) at 4th bOPV dose | 15.9 [15.4-16.9] | 16 [15-17] |
| Experienced delay in bOPV birth dose initiation (1st vaccination at >7 days old) | 39.8% (41) | 42.4% (28) |
| Received ≥ 1 IPV dose | 100% (103) | 100% (66) |
| Completed rotavirus Rotarix vaccine series | 100% (103) | 100% (66) |
| **Anthropometrics, nutrition and clinical history at the time of fourth bOPV dose** | | |
| Low birthweight (<2.5 kg) | 7.9% (8) | 6.3% (4) |
| Exclusively breastfed[c] | 91.3% (94) | 90.9% (60) |
| Stunting | 30.4% (31/102) | 30.3% (20) |
| HIV exposed uninfected | 8.7% (9) | 7.6% (5) |
| Any detected infection | | |
| HIV infected | 0.0% (0) | 0.0% (0) |
| Schistosome CAA positive (≥30 ug/ml) | 13.9% (14/101) | 16.9% (11/65) |
| *Giardia lamblia* | 4.9% (5) | 4.6% (3) |
| Other parasitic ova detected by Kato Katz[d] | 13.6% (14) | 16.7% (11) |
| Enteroinvasive bacterial pathogen associated with dysentery[e] | 11.7% (12) | 9.1% (6) |
| Other bacterial pathogens[f] | 70.9% (73) | 69.7% (46) |
| Viral pathogens[g] | 39.8% (41) | 34.9% (23) |
| High burden EE (composite score ≥7)[h] | 29.4% (30/102) | 31.8% (21) |
| Neopterin (NEO) concentration (ng/ml) | 0.35 [0.16-0.94], n=102 | 0.32 [0.16-0.94] |
| Myeloperoxidase (MPO) concentration (ng/ml) | 1855 [1394-4076], n=102 | 1994 [1385-4560] |
| Alpha-1-antitrypsin (AAT) concentration (ug/L) | 2248 [1112-5653], n=102 | 2393 [1124-6305] |

[a]The denominator is 103 for the full longitudinal cohort and 66 for the cohort with complete data unless otherwise specified.
[b]Calculated using the Oxford Multidimensional Poverty Index, which ranges from 0 to 1, with higher scores indicating greater levels of multidimensional poverty. Available at: Oxford Poverty & Human Development Initiative. The Global Multidimensional Poverty Index. https://ophi.org.uk/global-mpi
[c]For the longitudinal cohort, current breastfeeding status was reported by caregivers at baseline.
[d]Amoeba, *Ascaris* worm, Hookworm, Tapeworm, *Hymenolepis nana.*
[e]Enteroinvasive *Escherichia coli,* Shigella-toxin producing *E. coli, Campylobacter*, *Salmonella*.
[f]*Clostridium difficile, Plesiomonas shigelloides*, Enterotoxigenic *E. coli,* Enteropathogenic *E. coli,* Enteroaggregative *E. coli.*
[g]Rotavirus, adenovirus, sapovirus, astrovirus, norovirus.
[h]Composite EE score (0–10) calculated from NEO, MPO and AAT concentrations using the method described in Arndt, et al. 2016.[1] Composite EE scores in the upper quartile (≥7) were defined as high burden EE.
[1]Arndt, M.B., et al., Fecal Markers of Environmental Enteropathy and Subsequent Growth in Bangladeshi Children. Am J Trop Med Hyg, 2016. 95(3): p. 694-701.

**Supplementary Figure 3:** Longitudinal trajectory of type-1 (PV1) and type-3 poliovirus (PV3) specific $\log_2$ neutralisation titre and $\log_{10}$ IgA MFI in stool (purple) and serum (red) samples alongside $\log_{10}$ IgG MFI in serum (red) samples among the longitudinal sample including only participants with no missing outcome data due to missing visits or missing serum/stool neutralisation data (n=66), stratified by positive neutralisation in stool at baseline (>=1:8). The bolded line represents the medians at each visit, and the transparent band represents the interquartile range.

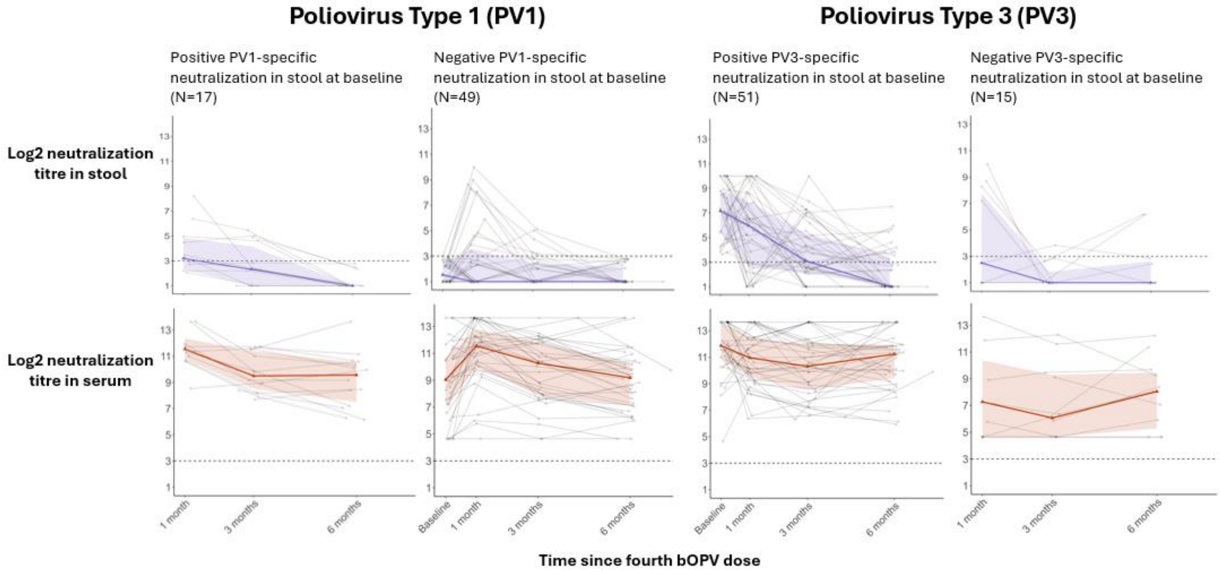

**Supplementary Figure 4:** Association between selected characteristics and recording a detectable (≥1:4) type-1 (PV1; red) and type-3 poliovirus (PV3; blue) neutralisation titre at any timepoint (between baseline and month-6) among the longitudinal sample (n=103), fitted using a logistic regression adjusted for participant age. The square represents the odds ratio, and the bar represents the 95% confidence interval (CI).

■ Detectable neutralization for poliovirus type-1 ■ Detectable neutralization for poliovirus type-3

| Variable | PV1 OR [95% Ci], p | PV3 OR [95% Ci], p |
|---|---|---|
| Non-exclusive breastfeeding[1] | 2.38 [0.54–10.56], p=0.255 | 1.27 [0.14–11.44], p=0.833 |
| At least 15 minutes to water-source | 1.13 [0.37–3.50], p=0.828 | 0.86 [0.20–3.63], p=0.836 |
| Stunting | 0.59 [0.20–1.74], p=0.342 | 1.19 [0.26–5.45], p=0.819 |
| HIV exposed uninfected | 0.82 [0.15–4.41], p=0.818 | 0.28 [0.05–1.63], p=0.157 |
| Schistosomiasis CAA | 1.47 [0.30–7.20], p=0.635 | NA[2] |
| Enteroinvasive bacteria[3] | 0.64 [0.16–2.64], p=0.537 | 0.47 [0.09–2.54], p=0.379 |
| Other bacteria[4] | 1.17 [0.40–3.46], p=0.770 | 0.98 [0.23–4.19], p=0.979 |
| Virus[5] | 0.67 [0.25–1.84], p=0.438 | 0.25 [0.06–1.02], p=0.054 |
| High burden environmental enteropathy[6] | 1.23 [0.40–3.78], p=0.721 | 0.21 [0.05–0.85], p=0.028 |

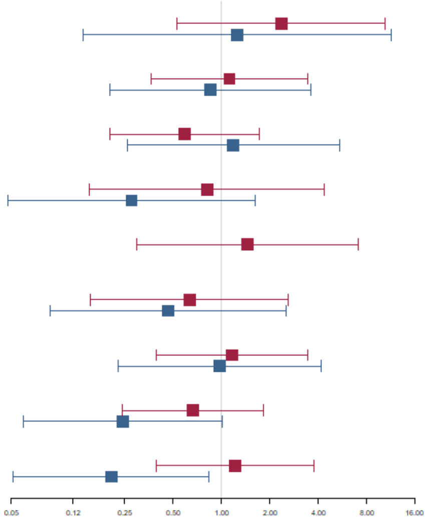

[1]Current breastfeeding status was reported by caregivers at baseline; [2]All participants who tested positive for schistosomiasis CAA also had a positive neutralisation response to type-3 poliovirus; [3]Includes: Enteroinvasive *E. coli*, Shigella-toxin producing *E. coli,* Campylobacter, Salmonella; [4]*Clostridium difficile, Plesiomonas shigelloides*, Enterotoxigenic *E. coli,* Enteropathogenic *E. coli,* Enteroaggregative *E. coli;* [5]rotavirus, adenovirus, sapovirus, astrovirus, norovirus. [6]High burden environmental enteropathy (EE) is defined as falling in the upper quartile of composite EE scores (i.e. ≥7).

**Supplementary Figure 5:** Distribution of poliovirus type-2 (PV2) specific $log_2$ neutralisation titres and $log_{10}$ IgA and IgG MFI in stool and serum samples after vaccination with four doses of bOPV plus one dose of IPV through the routine immunisation system, among participants in a longitudinal sample (orange points, n=343 observations among 103 participants) and a cross-sectional sample (blue points, n=246). Participants in the 8-9 year cohort received tOPV but did not receive IPV, through routine immunisation. The x-axis reflects time after fourth bOPV dose, with baseline measures collected during the week of the fourth dose. Horizontal lines indicate the median; box and whisker indicate the interquartile range (IQR) and 1.5*IQR. Time increments on the x-axis are not represented linearly.

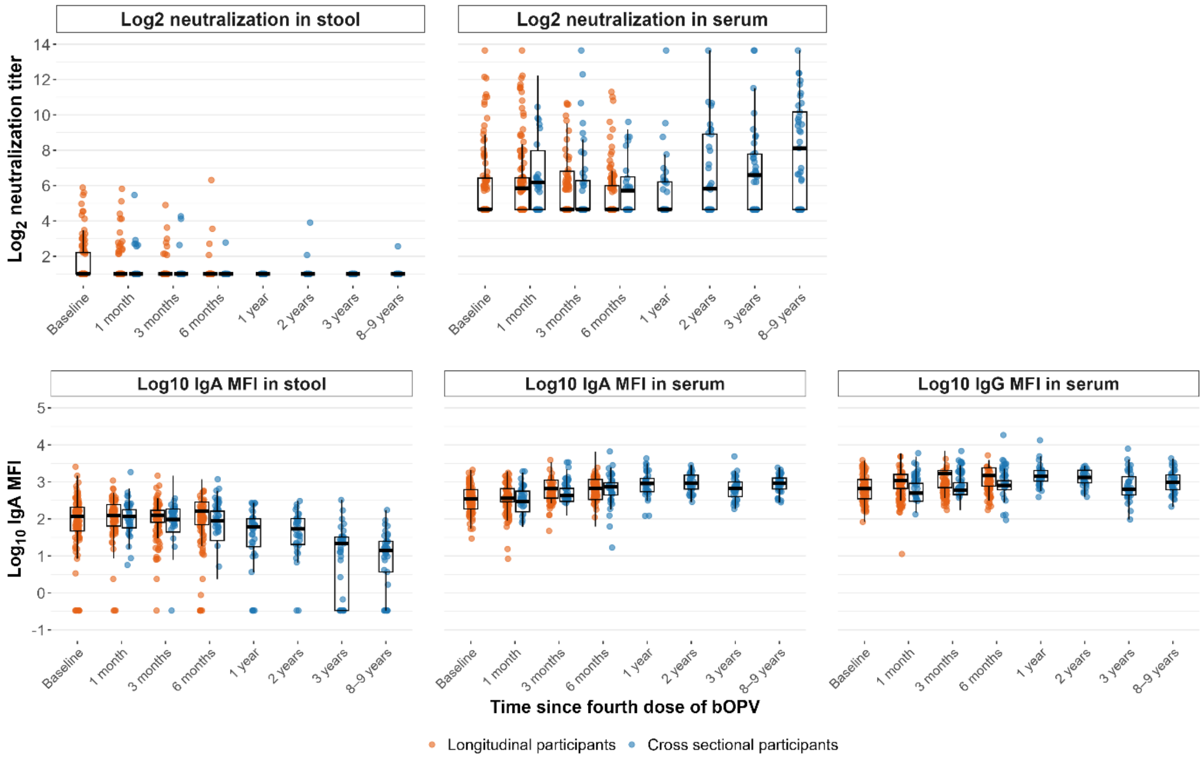
